# Discontinuation of Heart Failure Therapy in patients Undergoing Non-Cardiac surgery: Data from a Real-world Cohort

**DOI:** 10.1101/2023.05.05.23289601

**Authors:** Malik Elharram, Xiaoming Wang, Pishoy Gouda, Michelle M. Graham

## Abstract

**Background and Aims:** Patients with heart failure (HF) with reduced ejection fraction (HFrEF) are at high risk for cardiovascular events following non-cardiac surgery. The perioperative period represents many challenges to maintain guideline directed medical therapy (GDMT). We examined GDMT use in HFrEF patients following non-cardiac surgery, and the association of medication changes with cardiovascular outcomes.

**Methods:** Using linked administrative databases, a retrospective cohort of HFrEF patients undergoing major non-cardiac surgery between 2008 and 2020 was formed. Pre-operative use of GDMT was determined by outpatient prescriptions up to 90 days prior to surgery. Changes in GDMT was defined as discontinuation or a dose reduction (≥50%) of baseline therapies at 90 days after discharge. The primary composite outcome was HF hospitalization or all-cause mortality at one-year adjusted for age, sex, components of the Revised Cardiac Risk Index and the Charlson Comorbidity index.

**Results:** Of 397,829 index surgeries, there were 7667 (2%) patients with pre-existing HFrEF on at least one GDMT (50.6% female; mean age: 75 +/- 12 years). At 90 days post-operatively, 46% of patients had undergone major changes to GDMT. Compared to patients who continued GDMT, patients with any change to therapy had a higher incidence of the primary outcome (52% vs. 46%, aOR: 1.14, 95% CI: 1.03-1.25) and all-cause mortality at one year (8.5% vs. 4.9%, aOR: 1.57, 95% CI: 1.3-1.90).

**Conclusion:** Among patients with HFrEF undergoing major non-cardiac surgery, few are on optimal GDMT, and perioperative changes to GDMT is associated with higher odds for HF hospitalization or death.

## Introduction

Heart failure (HF) is highly prevalent among older adults undergoing non-cardiac surgery, and with a rising elderly population with underlying cardiovascular risk, this estimate is projected to continue to increase.^1^ HF is an established independent risk factor for adverse perioperative outcomes and is associated with a three to five-fold increase in perioperative mortality.^2,3^ Patients with HF with reduced ejection fraction (HFrEF) are a subset of patients at higher risk, and depressed left ventricular ejection fraction is an important predictor of adverse cardiovascular outcomes following surgery.^4^ Guideline directed medical therapy (GDMT) including a beta blocker (BB), renin angiotensin system inhibitor (RASi), mineralocorticoid receptor antagonist (MRA), and recently a sodium glucose co-transporter 2 inhibitor (SGLT-2i), improve morbidity and mortality in HFrEF,^5-10^ yet their utilization in the perioperative period remains unclear.

The pathogenesis of cardiovascular events in the perioperative period results from focal surgical or anesthetic triggers that initiate inflammatory, hypercoagulable, and hypoxic states,^11^ which may lead to decompensation in patients with pre-existing HFrEF. Furthermore, post-operative hypotension and renal failure may lead to challenges in reinitiating or continuing GDMT, which may worsen short and long-term HF outcomes. In prior studies of non-surgical hospitalized patients with HFrEF, discontinuation of RASi, BB or MRA at discharge was associated with an increased risk for HF readmission, and short and long-term mortality.^12-14^

The perioperative period represents many challenges to maintain guideline directed medical therapy (GDMT) in HFrEF patients following major surgery. We aim to examine the contemporary use of GDMT for HFrEF following non-cardiac surgery, and the association of changes to these therapies with HF related outcomes.

## Methods

### Data Source

A population-based cohort of patients who underwent non-cardiac surgery in the province of Alberta, Canada was formed, which has previously been described.^15^ The dataset was created by linking 5 administrative databases in Alberta using each patients’ provincial health number: (1) Alberta Inpatient Discharge Abstract Database, which includes information on all admissions to acute care facilities including admission diagnosis and inpatient surgical procedures; (2) the Pharmaceutical Information Network (PIN) database, which includes records of every individual’s active and previous prescription outpatient medications in Alberta, regardless of coverage. Over the counter medications are not captured in this database; (3) the Alberta Health Care Practitioner Claims Database, which includes physician claims for outpatient services; (4) Alberta Registry data, in which deaths are recorded; and (5) the Alberta Health Services Laboratory database, which is the repository for all inpatient and outpatient laboratory investigations.

### Population selection

We identified a cohort of surgical patients with pre-existing HFrEF who were discharged alive following hospitalization for major non-cardiac surgery in Alberta. We examined patients admitted for major surgical procedures pertaining to intrathoracic, orthopaedic, intraperitoneal, vascular, and pelvic surgeries **(Supplementary Table 1)** If an individual underwent >1 surgical procedure during the study period, only the first surgery was used. Patients needed to have filled a prescription for either a RASi (angiotensin converting enzyme inhibitor, angiotensin receptor blocker, or angiotensin receptor-neprilysin inhibitor), BB, or MRA within 90 days (to account for stockpile medications) prior to surgery. We did not consider the use of SGLT-2i as our cohort preceded the widespread adoption of these medications into HF guidelines. Surgical procedures were identified using ICD-9-CM and CCI procedural codes, as per our prior study.^15^ We excluded patients undergoing cardiac or minor surgical procedures. Patients with HFrEF were defined as having at least 1 inpatient admission or at least 2 outpatient clinic visits with diagnosis of HFrEF by ICD code within 5 years of surgery (*ICD-9-CM* or *ICD-10-CM*) codes 428.2 or I50.2 (systolic HF) or 428.4 or I50.4 (combined systolic and diastolic HF).^16^

### Exposure

The exposure group was based on continuation of preoperative doses of GDMT (BB, RASi, and/or MRA) compared to changes to therapy, which were defined as either discontinuation or dose reduction of therapy. Baseline use of GDMT was defined as a medication that was dispensed within 90 days prior to admission for surgery to account for stockpiled medication. Continuation status was defined as patients having a record of an outpatient prescription filled within 90 days following discharge from non-cardiac surgery. 90-days was used as our threshold as that is the longest interval that a pharmacy can dispense medications in our jurisdiction. Our secondary exposure was a dose reduction of HF therapy, defined as a ≥50% reduction in dose from baseline, within 90 days following discharge. Data on medication use were extracted from the pharmaceutical information network (PIN) database.

### Covariates

Baseline comorbidities were obtained from the discharge abstract database using International Classification of Diseases, Tenth Revision codes (**Supplementary Table 2)**. This included components of the Revised Cardiac Risk Index (RCRI) score ^17^ (1) history of ischemic heart disease; (2) heart failure; (3) stroke or transient ischemic attack; (4) insulin-treated diabetes; (5) creatinine ≥ 177 μmol/L (from most recent creatinine in the Alberta Health Services Laboratory database); and (6) high-risk surgery (intrathoracic, vascular, and intraperitoneal). Pre-operative BNP and post-operative creatinine were further obtained from the Alberta Health Services Laboratory database. We also included any prior history of atrial fibrillation, type 1 or type 2 diabetes mellitus, hypertension, dyslipidemia, and components of the Charlson comorbidity index.^18^

### Outcomes

Our primary outcome was a composite of all-cause mortality or hospitalization for HF at one year. HF hospitalization was defined as an inpatient admission for HF using a hospital inpatient record using ICD-10 codes (I50, I110). Vital status was obtained from the Alberta registry. Patients were followed from the date of cohort entry (90 days after discharge from non-cardiac surgery) until the occurrence of the outcome of interest or until the end of follow-up at one year.

### Statistical analysis

Descriptive analysis compared baseline characteristics of patients who continued GDMT without any changes, compared to patients who underwent changes (either discontinuation or dose reduction) using mean and standard deviation (SD) for continuous variables and frequency distribution for categoric variables. We stratified changes in GDMT based on their baseline use of either one, two or all three GDMT prior to non-cardiac surgery. A logistic regression model was used to examine predictors of changes in GDMT. We estimated an adjusted odds ratio with 95% CI to assess the one-year odds for readmission for HF or all-cause mortality, adjusted for: age, sex, components of RCRI risk score (history of ischemic heart disease, stroke or TIA, insulin treated diabetes, AKI (Cr >177) and high-risk surgery) and Charlson comorbidity index. SAS Enterprise Guide 8.3 (SAS Institute) was used for all statistical analyses.

## Results

### Baseline characteristics

From October 1^st^, 2008 - September 30^th^, 2020, there were 552,224 surgeries performed in the province of Alberta, which included 397,829 major non-cardiac surgical procedures. Of those, we identified 7667 (1.9%) patients with a diagnosis of HFrEF who were on at least one class of GDMT prior to surgery (RASi, BB, or MRA) **(Supplementary Figure 1)**. The mean age of patients were 75 years +/-12.3 years and 51.7% were female. The most common surgical procedures performed were orthopaedic in 4067 (53%) followed by intraperitoneal 2475 (32%) and pelvic 699 (9%). Following major non-cardiac surgery, 3546 patients (46%) experienced changes to baseline GDMT, which included discontinuation of at least one class of medication in 2334 (30%) or dose reduction >50% in a further 1212 (16%) patients. Individuals who had there HF therapy discontinued or dose reduced were more likely to be older, female, and have a higher prevalence of chronic kidney disease, cerebral vascular disease, atrial fibrillation/flutter, or HF hospitalization within the prior year. Pre and post operative creatinine and pre-operative BNP (where available) were higher amongst patients who underwent changes to GDMT (**Table 1)**.

**Table 1.** Baseline Clinical Characteristics.

|  | Overall cohort<br>(n= 7667) |  | No<br>discontinuation/reduction<br>in therapy (n=4121) |  | Discontinuation or<br>reduction in therapy<br>(n=3546) |  | p-value |
| --- | --- | --- | --- | --- | --- | --- | --- |
| Age (Median, IQR) | 7667 | 77 (67 - 84) | 4121 | 75 (66 -83) | 3546 | 78 (69 -86) | <.0001 |
| Sex (% Female) | 7667 | 3963<br>(51.69%) | 4121 | 1926 (46.74%) | 3546 | 1778 (50.14%) | 0.0029 |
| **Past Medical History** |  |  |  |  |  |  |  |
| Hypertension (%) | 7667 | 5772<br>(75.28%) | 4121 | 3092 (75.03%) | 3546 | 2680 (75.58%) | 0.5793 |
| Dyslipidemia (%) | 7667 | 1991<br>(25.97%) | 4121 | 1081 (26.23) | 3546 | 910 (25.66%) | 0.5712 |
| Insulin dependent Type 2 Diabetes Mellitus (%) | 7667 | 1090<br>(14,22%) | 4121 | 582 (14.12%) | 3546 | 508 (14.33%) | 0.7995 |
| Non insulin dependent Type 2 Diabetes Mellitus (%) | 7667 | 2086<br>(27.21%) | 4121 | 1127 (27.35%) | 3546 | 959 (27.04%) | 0.7662 |
| Ischemic heart disease (%) | 7667 | 3373<br>(43.99%) | 4121 | 1794 (43.53%) | 3546 | 1579 (44.53%) | 0.3811 |
| Peripheral vascular disease (%) | 7667 | 1055<br>(13.76%) | 4121 | 559 (13.56%) | 3546 | 496 (13.99%) | 0.5920 |
| Cerebral vascular disease (%) | 7667 | 856<br>(11.16%) | 4121 | 432 (10.48%) | 3546 | 424 (11.96%) | 0.0410 |
| Chronic Kidney Disease (%) | 7667 | 1282<br>(16.72%) | 4121 | 633 (15.36%) | 3546 | 649 (18.30%) | 0.0006 |
| Atrial fibrillation/flutter (%) | 7667 | 3166<br>(41.29%) | 4121 | 1567 (38.02%) | 3546 | 1599 (45.09%) | <.0001 |
| History of HF admission within the past year (%) | 7667 | 5225<br>(68.15%) | 4121 | 2674 (64.89%) | 3546 | 2551 (71.94%) | <.0001 |
| Charlson comorbidity index (mean, SD) | 7667 | 1.39<br>(1.20) | 4121 | 1.26 (1.14) | 3546 | 1.54 (1.25) | <.0001 |
| RCRI score (mean, SD) | 7667 | 1.89<br>(1.19) | 4121 | 1.85 (1.20) | 3546 | 1.94 (1.18) | 0.0016 |
| **Surgery type** |  |  |  |  |  |  |  |
| Vascular (%) | 7667 | 316<br>(4,12%) | 4121 | 183 (4.44%) | 3546 | 133 (3.75%) | 0.1297 |
| Intraperitoneal (%) | 7667 | 2475<br>(32.28%) | 4121 | 1410 (34.21%) | 3546 | 1065 (30.03%) | <.0001 |
| Intrathoracic (%) | 7667 | 110<br>(1.43%) | 4121 | 71 (1.72%) | 3546 | 39 (1.10%) | 0.0222 |
| Pelvic (%) | 7667 | 699<br>(9.12%) | 4121 | 438 (10.63%) | 3546 | 261 (7.36%) | <.0001 |
| Orthopedic (%) | 7667 | 4067<br>(53.05%) | 4121 | 2019 (48.99%) | 3546 | 2048 (57.76%) | <.0001 |
| **Laboratory investigations** |  |  |  |  |  |  |  |
| Pre-operative Creatinine (median, umol/L) | 5264 | 99 (78 – 135) | 2763 | 97 (77 – 130) | 2501 | 101 (78 – 140) | 0.0017 |
| Pre-operative BNP (median, ng/ml) | 676 | 403<br>(188.5 – 781.5) | 299 | 357 (174 – 670) | 377 | 434 (198 - 878) | 0.0178 |
| Pre-operative NT-proBNP (median, ng/ml) | 540 | 1908<br>(652 – 4531.5) | 248 | 1640.5 (568.5 – 4037) | 256 | 2102 (728 – 4752.5) | 0.0763 |
| Post-operative creatinine (median, | 5838 | 95 (74 – 130) | 3044 | 94 (73 – 127) | 2794 | 97 (75 – 135) | 0.0054 |
| μmol/L) |  |  |  |  |  |  |  |

**Legend:** SD: standard deviation, IQR: Interquartile range, HF: Heart failure, BNP: Brain natriuretic peptide,

### Guideline Directed Medical Therapy Utilization

Overall, 5724 (74%) of patients with HFrEF were on monotherapy with GDMT prior to non-cardiac surgery. For patients on monotherapy, the most common GDMT class was a RASi (87%), followed by an MRA (8%), and a beta blocker (5%). Two agents were used in 1631 (21%) of patients prior to non-cardiac surgery, with the most common combination being a RASi+ MRA (54%), followed by a BB + RASi (41%). Only 312 (4%) of patients were on triple therapy **(Table 2 & Supplementary Table 3)**. At 90 days following major non-cardiac surgery, 4131 (72.2%) of patients on one, 1019 (62.5%) on two, and 183 (58.7%) on all three GDMT classes continued their medications without changes. For patients on monotherapy, an MRA was the most frequently discontinued therapy after surgery in 150 patients (34%) of prior users. For patients on dual therapy, a BB + MRA and a RASi + MRA were more likely to have discontinued at least one medication during follow up. For patients on triple therapy at baseline, 129 (42%) of patients discontinued at least one therapy.

**Table 2.** Discontinuation of Guideline Directed Medical Therapy Following Major Non-Cardiac Surgery.

|  | Pre-operative use of GDMT (n= 7667) | 90-days following non-cardiac surgery |  |  |  |
| --- | --- | --- | --- | --- | --- |
|  |  | Continued all therapies (N= 5333) | Discontinuation of 1 class (N= 1991) | Discontinuation of 2 classes (N= 304) | Discontinuation of 3 classes (N= 39) |
| On one therapy | 5724 (74.66%) | 4131 (72.17%) | 1593 (27.83%) |  |  |
| On two therapies | 1631 (21.27%) | 1019 (62.48%) | 333 (20.42%) | 279 (17.11%) |  |
| On three therapies | 312 (4.07%) | 183 (58.65%) | 65 (20.83%) | 25 (8.01%) | 39 (12.50%) |

**Legend:** GDMT: Guideline directed medical therapy; One therapy: Either beta blocker (BB) or renin angiotensin system inhibitor (RASi), or mineralocorticoid receptor blocker (MRA); On two therapies: combination of RASi+ MRA, BB+ RASi, or MRA+BB; On three therapies: BB+MRA+RASi

Dose reduction of HF therapy occurred in 906 (22%) on one therapy, 264 (26%) on two therapies, and 42 (22%) of patients on three therapies. For patients on monotherapy, the most common medication to undergo a dose reduction was an MRA in 68 patients (24%). For patients on dual therapy the most common combination to undergo a dose reduction of at least one therapy was a BB+ MRA (28%) followed by a RASi+ MRA (26%) (**Table 3 & Supplementary Table 4)**. Factors associated with discontinuation or dose reduction to GDMT therapy at follow-up included older age, prior history of atrial fibrillation/flutter, heart failure admission in the prior year, orthopaedic surgery, and elevation in preoperative BNP and post operative creatinine **(Table 4)**.

**Table 3.**
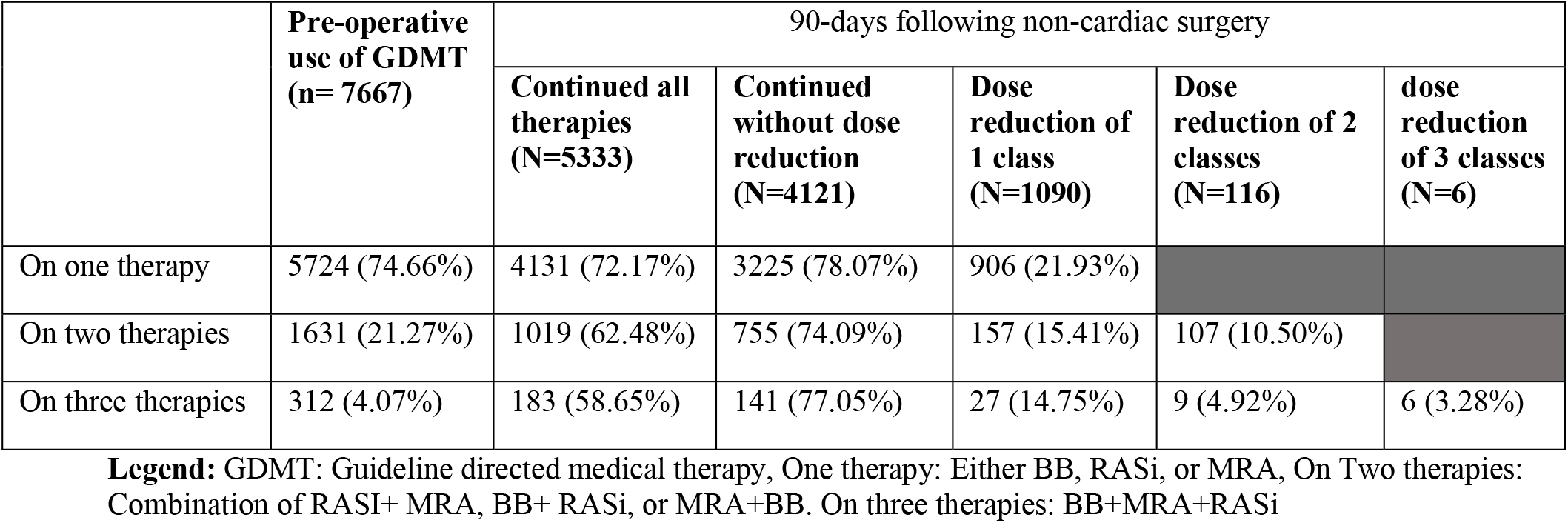
Dose Reduction of Guideline Directed Medical Therapy Following Major Non-Cardiac Surgery.

|  | Pre-operative use of GDMT (n= 7667) | 90-days following non-cardiac surgery |  |  |  |  |
| --- | --- | --- | --- | --- | --- | --- |
|  |  | Continued all therapies (N=5333) | Continued without dose reduction (N=4121) | Dose reduction of 1 class (N=1090) | Dose reduction of 2 classes (N=116) | dose reduction of 3 classes (N=6) |
| On one therapy | 5724 (74.66%) | 4131 (72.17%) | 3225 (78.07%) | 906 (21.93%) |  |  |
| On two therapies | 1631 (21.27%) | 1019 (62.48%) | 755 (74.09%) | 157 (15.41%) | 107 (10.50%) |  |
| On three therapies | 312 (4.07%) | 183 (58.65%) | 141 (77.05%) | 27 (14.75%) | 9 (4.92%) | 6 (3.28%) |

**Legend:** GDMT: Guideline directed medical therapy, One therapy: Either BB, RASi, or MRA, On Two therapies: Combination of RASi+ MRA, BB+ RASi, or MRA+BB. On three therapies: BB+MRA+RASi

**Table 4.**
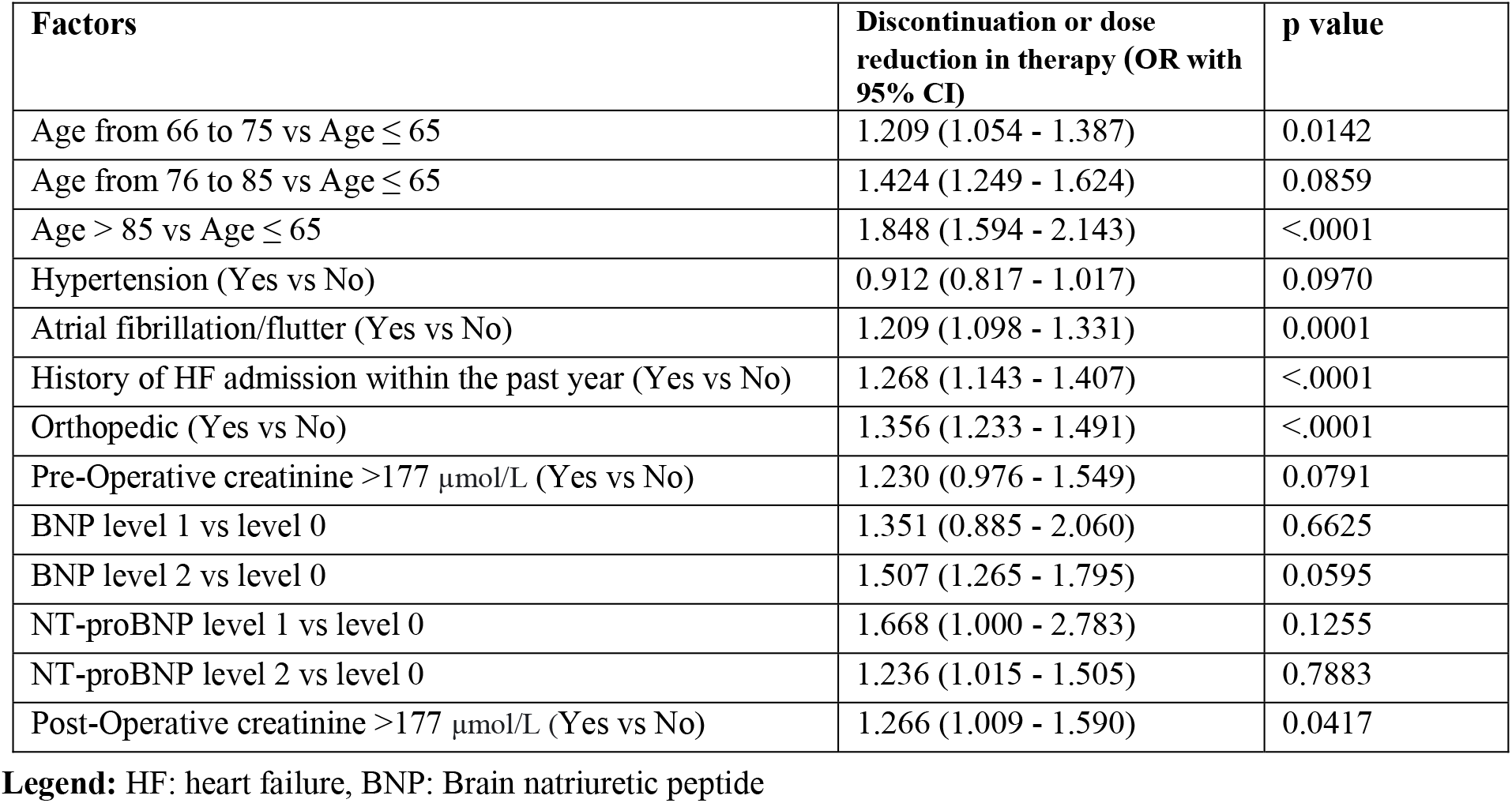
Factors associated with discontinuation or dose reduction in therapy following non-cardiac surgery.

| Factors | Discontinuation or dose reduction in therapy (OR with 95% CI) | p value |
| --- | --- | --- |
| Age from 66 to 75 vs Age ≤ 65 | 1.209 (1.054 - 1.387) | 0.0142 |
| Age from 76 to 85 vs Age ≤ 65 | 1.424 (1.249 - 1.624) | 0.0859 |
| Age > 85 vs Age ≤ 65 | 1.848 (1.594 - 2.143) | <.0001 |
| Hypertension (Yes vs No) | 0.912 (0.817 - 1.017) | 0.0970 |
| Atrial fibrillation/flutter (Yes vs No) | 1.209 (1.098 - 1.331) | 0.0001 |
| History of HF admission within the past year (Yes vs No) | 1.268 (1.143 - 1.407) | <.0001 |
| Orthopedic (Yes vs No) | 1.356 (1.233 - 1.491) | <.0001 |
| Pre-Operative creatinine >177 μmol/L (Yes vs No) | 1.230 (0.976 - 1.549) | 0.0791 |
| BNP level 1 vs level 0 | 1.351 (0.885 - 2.060) | 0.6625 |
| BNP level 2 vs level 0 | 1.507 (1.265 - 1.795) | 0.0595 |
| NT-proBNP level 1 vs level 0 | 1.668 (1.000 - 2.783) | 0.1255 |
| NT-proBNP level 2 vs level 0 | 1.236 (1.015 - 1.505) | 0.7883 |
| Post-Operative creatinine >177 μmol/L (Yes vs No) | 1.266 (1.009 - 1.590) | 0.0417 |

**Legend:** HF: heart failure, BNP: Brain natriuretic peptide

### Clinical Events

At one year follow-up, the composite of either HF hospitalization or death occurred in 3782 (49.3%) of patients. Compared to patients who continued GDMT, patients with any change to therapy had a higher incidence of the primary outcome (52% vs. 47%, aOR: 1.14, 95% CI: 1.04-1.25). Medication changes to patients on one and three therapies were associated with an increased odds for the primary outcome (one therapy: 52% vs. 48%, aOR: 1.13 (1.01-1.25); three therapies: 56% vs. 36%, aOR: 1.9, 95% CI: 1.17-3.1). Medication changes to patients on two therapies at baseline was not associated with an increased odds for the primary outcome (aOR: 1.07, 95% CI: 0.88-1.31) (**Table 5)**. Overall, 504 (7%) of patients died within one year following non-cardiac surgery. Compared to patients who continued their GDMT, patients with any change to GDMT had significant risk of mortality (8.5% vs. 4.9%, aOR: 1.57, 95% CI: 1.30-1.90).

**Table 5.** Discontinuation or Dose Reduction in Therapy on HF Hospitalization or All-cause Mortality at 1-year.

|  | Heart Failure Hospitalization or All-Cause Mortality |  |  |  |
| --- | --- | --- | --- | --- |
|  | No discontinuation or dose reduction in therapy | Discontinuation or dose reduction in therapy |  |  |
|  | Events n = (%) | Events n = (%) | Unadjusted OR (95% CI) | Adjusted OR* (95% CI) |
| On HF therapy at baseline (one, two or three therapies) | 1947 (47.25%) among 4121 | 1835 (51.75%) among 3546 | 1.197 (1.094, 1.310) | 1.140 (1.039, 1.251) |
| One therapy | 1533 (47.53%) among 3225 | 1292 (51.70%) among 2499 | 1.181 (1.064, 1.312) | 1.126 (1.010, 1.254) |
| Two therapies | 363 (48.08%) among 755 | 448 (51.14%) among 876 | 1.130 (0.930, 1.373) | 1.073 (0.879, 1.311) |
| Three therapies | 51 (36.17%) among 141 | 95 (55.56%) among 171 | 2.206 (1.396, 3.484) | 1.900 (1.176, 3.069) |

4 **Legend:** GDMT: Guideline directed medical therapy, HF: Heart failure, OR: Odds ratio, CI: Confidence Interval 5 **\*Adjusted odds ratio:** age, sex, components of revised cardiac risk index risk score and Charlson comorbidity 6 index.

Mortality was significantly higher in patients who underwent changes to one therapy (aOR: 1.55, 95% CI: 1.24-1.93), but without any significant difference in patients on two and three therapies, albeit a low number of events (**Supplementary Table 5)**.

## Discussion

In this large retrospective cohort of patients with HFrEF undergoing major non-cardiac surgery we observed that patients with HFrEF demonstrate a high rate of major cardiovascular events, with one in two being admitted for a heart failure exacerbation or death. Few patients with HFrEF are on optimal GDMT prior to surgery (4%) and nearly half of patients underwent significant dose reductions/discontinuation of GDMT in the peri-operative period. These changes to GDMT were associated with higher odds for HF hospitalization or mortality at one year.

Together, our findings identify important real-world gaps in GDMT utilization in HFrEF patients following major surgery and emphasize the need for careful perioperative assessment and monitoring, and close outpatient follow-up to improve treatment in line with current guideline recommendations.^19-21^

Pre-existing HF is among the most significant risk factors for perioperative cardiovascular complications. In an analysis of 23,340 chronic heart failure patients undergoing major non-cardiac surgery, the risk adjusted mortality within 30 days of surgery was 11.7% for patients with pre-existing HF compared to 6.6% for patients with coronary artery disease.^22^ Furthermore, there is a graded risk in patients with a lower left ventricular ejection fraction (EF). In a cohort of patients with chronic HF undergoing non-cardiac surgery, the 90-day postoperative mortality rate was lowest amongst patients with preserved EF (4.88%) and highest in patients with an EF of 30-39% (6.58%) and an EF of <30 % (8.34%).^23^ The elevated risk for poor outcomes extends beyond the perioperative period, with a cohort of patients with chronic HF undergoing elective or emergency surgery in Sweden demonstrating a one-year mortality of 16% for elective surgery and 39% for emergency surgery.^24^ Consistent with prior studies, we found that nearly half of patients with HFrEF undergoing major non-cardiac surgery were hospitalized for HF or died within the year following surgery. The long-term risk of cardiovascular events following surgery was comparable to data from patients admitted with a HF exacerbation,^25,26^ which remains one of the poorest prognostic factors in HF patients.^27^ Admission for major surgery in patients with HFrEF may signify an important clinical event akin to a HF hospitalization that requires careful clinical assessment and diligent outpatient follow-up.

While evidence from several landmark clinical trials have demonstrated the effectiveness of pharmacological therapy in improving morbidity and mortality in patients HFrEF,^5-10^ we noted a low proportion of patients on adequate (triple therapy) GDMT prior to non-cardiac surgery. Our findings appear to be consistent with studies done in the real-world setting, where GDMT in patients with HFrEF has been shown to be lower than levels achieved in clinical trials. In a retrospective observational study of 17,106 patients with HFrEF with an incident HF hospitalization, 23% of patients were not on any HF therapy on admission and only 13% were on triple therapy.^13^ Similarly in a large Canadian cohort of HFrEF, only 20% of patients were on triple therapy at 6 months following an index hospitalization for HF, and only 8.3% achieved optimal GDMT dose.^28^ Utilization of GDMT within our cohort was lower than these observational studies (only 4% on triple therapy), and may be related to an older cohort with more comorbidities, and more advanced HF. Additionally, roughly one third of patients in our cohort did not have a recent HF hospitalization, which is an important triggering event for medication assessment and titration.^29^ While current perioperative guidelines recommend adequate optimization of GDMT in HFrEF prior to elective major non-cardiac surgery,^30^ our findings from a generalizable non-cardiac surgical cohort suggest underperformance in the real-world setting.

We also found that significant changes to GDMT occurred in HFrEF patients following non-cardiac surgery. In our cohort, nearly one-half of patients experienced discontinuation or dose-reduction of therapy at 90 days following discharge. Hemodynamic or biochemical changes in the perioperative period may provide barriers to restarting or initiating GDMT in this population. Perioperative acute kidney injury (AKI) occurs between 6.3-13.4% of patients undergoing major non-cardiac surgery,^31^ and the presence of pre-existing HF further increases this risk.^32^ Additionally, multiple perioperative factors may lead to disruption of potassium homeostasis leading to hyperkalemia.^33^ Renal dysfunction and hyperkalemia in the perioperative setting may lead to hesitancy to restarting a RASi or an MRA. Indeed, within our cohort, an MRA or RASi were the most frequent medications to undergo changes or discontinuation, and pre-existing chronic kidney disease and postoperative creatinine were among the strongest predictors for changes in therapy. Furthermore, hypotension from the effects of anesthetic and analgesic agents, shifts in fluid balance, or bleeding may further serve as a barrier to continuing or restarting therapy. Our analysis also identified that age, renal dysfunction, underlying atrial fibrillation, and more advanced HF (higher BNP and recent HF admission) were associated with changes to GDMT. Under-titration and discontinuation of GDMT are known to occur more frequently in elderly patients with increased medical comorbidities,^34^ despite the robust benefit demonstrated in this population.^35^ This further illustrates the risk-treatment paradox in HF, where patients with the greatest need for GDMT are paradoxically the least likely to receive appropriate therapy.

Discontinuation or dose reduction of GDMT in HFrEF patients following surgery was associated with adverse outcomes, including a 14% increased odds for HF hospitalization or mortality at one year. This association appeared to be strongest in the subgroup of patients who were preoperatively treated with triple therapy, where HFrEF patients have been shown to achieve the most clinical benefit.^36^ Furthermore, we found that changes in GDMT were associated with a 57% increase in all-cause mortality at one year. This is consistent with prior observational studies suggesting worsening clinical outcomes in HF patients who discontinued GDMT following admission for HF.^12-14^ In a large observational study of patients admitted with decompensated HF, discontinuation of GDMT, including ACE inhibitors/ARBs, beta-blockers, and MRAs, were associated with a 30% increase in 1-year all-cause mortality compared with maintaining GDMT.^37^ Similarly in an analysis from the Get With The Guidelines-Heart Failure Registry, discontinuation of ACEi/ARB among patients hospitalized for HFrEF was associated with a significantly higher 90-day and 1-year mortality and 30-day HF readmission compared to those who continued therapy.^14^ Finally, doses of GDMT below the target recommended by clinical trials ^5-10^ have been associated with poorer outcomes.^38^ Our results extend these prior observations to a non-cardiac surgical HFrEF population, and further illustrate the harm that discontinuation or dose reduction of GDMT may have on long-term clinical outcomes.

Our findings demonstrate important limitations in the contemporary management of HFrEF patients undergoing major non-cardiac surgery and emphasize the need for a coordinated and multidisciplinary approach to care and follow-up of this high-risk cohort. Following surgery, patients with HFrEF may benefit from a multidisciplinary care model between surgery, anesthesia, and cardiology/internal medicine to ensure adequate clinical assessment of hemodynamics, volume status, and biochemistry to target early re-initiation of GDMT. The perioperative setting may further serve as a key opportunity to address gaps in medical therapy including initiation, titration or switching of therapies prior to discharge. It remains imperative that HFrEF patients have early follow-up with an outpatient specialist for reassessment of clinical status and consideration for early medication reinitiation/titration, to improve treatment in keeping with guideline recommendations.^19-21^

## Limitations

The present results should be taken in the context of several limitations. As this was an administrative dataset, we were unable to capture echocardiographic measures of EF or functional class as markers of more advanced HF. However, the use of ICD-10 codes for patients with HFrEF, has been recently shown to accurately identify patients with reduced ejection fraction when defined as an EF <50%.^39^ Furthermore, we were unable to capture the rationale for discontinuation or dose reduction to therapy. Although we attempted to adjust for clinically important comorbidities in the relationship between changes in therapy and outcomes, the possibility for residual and unmeasured confounding exists. Finally given the period of study (2008-2020) our cohort predates the widespread use of SGLT-2i into HF guidelines, and we therefore did not consider the use these classes of medications in our study.

## Conclusion

In conclusion, among a generalizable cohort of patients with HFrEF undergoing major non-cardiac surgery, few (4%) are on optimal GDMT pre-operatively. Furthermore roughly half of patients with HFrEF will undergo further significant reductions in their GDMT post-operatively. These deleterious perioperative changes to GDMT are associated a 14% increased risk of HF hospitalization and all-cause mortality. Our findings confirm the need for careful multidisciplinary perioperative assessment and early outpatient follow-up to improve treatment and outcomes in this high-risk population.

## Data Availability

Data not available - participant consent

